# An efficient UV-C disinfection approach and biological assessment strategy for microphones

**DOI:** 10.1101/2022.07.09.22277021

**Authors:** Valentina Vignali, Tobi Hoff, Jacqueline de Vries-Idema, Anke Huckriede, Jan Maarten van Dijl, Patrick van Rijn

## Abstract

Hygiene is a basic necessity to prevent infections and though it is regarded as vital in general, its importance has been stressed again during the pandemic. Microbes may spread through touch and aerosols and thereby find their way from host to host. Cleaning and disinfection of possibly contaminated surfaces prevents microbial spread thus reducing potential illnesses. One item that is used by several people in a way that promotes close contact by touch and aerosol formation is the microphone. A microphone is a complex piece of equipment with respect to shape and various materials used to fabricate it and, hence, its disinfection is challenging. A new device has been developed to efficiently sterilize microphones using UV-C and the biological assessment has been done to identify its efficacy and translatability. For this investigation, a contamination procedure was developed using M13 bacteriophage as a model to illustrate the effectiveness of the disinfection. The susceptibility to UV-C irradiation of M13 in solution was compared to that of PR8 H1N1 influenza virus, which has a similar UV-C susceptibility as SARS-CoV-2. It was found that 10 min of UV-C treatment reduced the percentage of infectious M13 by 99.3% based on whole microphone inoculation and disinfection. UV-C susceptibility of M13 and influenza in suspension were found to be very similar, indicating that the microphone sterilization method and device function are highly useful and broadly applicable.

## 1. Introduction

Pandemics and antimicrobial resistance represent serious threats to global health [1,2]. Infections are an eminent and urgent problem within the medical field causing also many medical implant-related complications for which new treatment approaches are being developed [3,4]. While a lot of focus is on research for novel therapeutics, an increasing effort is being placed on prevention, since treatments may sometimes be unavailable or ineffective. Viruses are a well-known source of infection, which has become very apparent since the pandemic associated to COVID-19 [5,6] making an impact beyond comprehension [7,8][9]. Therefore, methods that enhance hygiene in all aspects of life are deemed critical to maintain a healthy lifestyle both personally and societally. In light of these eminent remaining threats, proper disinfection becomes a pertinent approach to subdue the problem [10]. Preventing the transmission of microbes through objects used by various individuals reduces the risk of infections. Disinfection of such objects could refrain the microbes from spreading and thereby prevent unnecessary suffering. One such item is a microphone.

A microphone is typically a device that is shared by various individuals and as the microphone is in close contact with the oral cavity and aerosols, the chance for microbes to transfer from the oral cavity to the surface of the microphone is high (**Figure 1**). Hence, the industry that uses microphones would benefit substantially from a procedure that enables disinfection of microphones and thereby lowers the chances of possible spread and transmission of microbes, including SARS-CoV-2.

**Figure 1.**
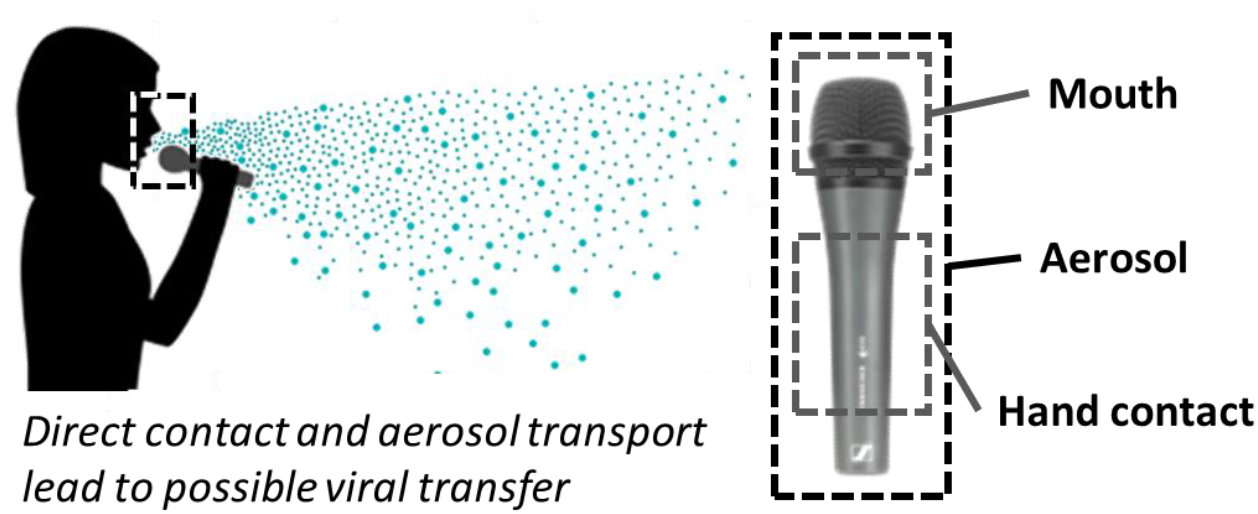
Representation of possible aerosol production during sound emission, and possible microbial contamination of different parts of a microphone, which may lead to the transmission of pathogens upon shared use of the same device by different individuals.

Here a new device has been tested that would allow disinfection of microphones by using UV-C irradiation. UV-C is known to damage viruses and bacteria rendering them non-infectious by directly damaging their genetic material (DNA/RNA) and proteins [11]. Other approaches for disinfection are known, involving the application of alcohol, hydrogen peroxide, or autoclaving, but these methods could damage the microphones. Disinfection with UV-C is an effective approach and, when applied in a short-enough timespan, it will not harm the material in question. It is therefore regarded as a useful disinfection tool in hospitals and more general settings to prevent the transmission of SARS-CoV-2 [12,13]. The use of UV-C has already been applied to inactivate various microorganisms [14,15], especially bacteria [16,17] and viruses [11,18,19], including SARS-CoV-2 [20–22], in different conditions, such as aerosols [19], surfaces [23,24], medical materials [25], dairy products [26], and other foods [27]. Accordingly, we developed a novel UV-C irradiation device for the disinfection of microphones. To test the efficacy of our device, microphones were inoculated/contaminated with a model virus, the M13 bacteriophage. The M13 bacteriophage was used as a representative model to be able to test the device and our experimental protocol in a biologically safe manner, as this virus is non-infective towards humans [28,29]. To illustrate the general applicability of the device, UV-C treatment of M13 in suspension was compared to UV-C treatment of influenza virus in suspension, which was previously shown to respond to UV-C in a similar manner as SARS-CoV-2 on contaminated solid surfaces [30].

## 2. Materials and Methods

### M13 bacteriophage

The M13 bacteriophage used in the experiments was isolated from *E. coli* ER2738/M13KE gIII following a protocol described in literature [31]. Bacterial strain: *E. coli* K12 ER2738 (New England Bio Labs E4104S) [Genotype: F’ *proA*+*B*+ *lacI*q Δ(lacZ)M15 zzf::Tn10(TetR)/*fhuA2* ĝlnV Δ(*lac*-*proAB*) *thi*-1 Δ(*hsdS*-*mcrB*)5]. The M13KE gIII vector was derived from the cloning vector M13mp19, which carries the *lacZ*α gene (New England BioLabs).

### M13 bacteriophage titration

The concentration of the M13 bacteriophage was determined via the method of viral titration. 10-20 ml of LB were inoculated with *E. coli* K12 ER2738 isolated from a single colony cultured on a Lysogeny Broth (LB) agar plate supplemented with …μg/ml tetracycline and the resulting culture was incubated in a shaking incubator until the mid-log phase (OD_600_ ∼ 0.5). Series of 10-fold dilutions of the phage samples were prepared in duplicate. 10 μl of the dilution samples were transferred to a 100 μl aliquot of *E. coli* K12 ER2738 mid-log phase. The samples were vortexed and transferred to a static incubator at 37°C for 5-10 min. Subsequently, the samples were plated on LB Agar supplemented with … μg/ml IPTG and μg/ml Xgal and incubated over night at 37°C. Blue plaques were counted and used to determine the plaque-forming units (PFU) of the original sample. The preparation of medium and agar plates were done according to protocols that were previously described [31].

### Influenza virus

For the comparison between M13 and influenza, the A/Puerto Rico /8/ 34 (PR8) H1N1 influenza virus strain was used. This strain and the respective titration method that we applied were previously described [32].

### Comparison of M13 and influenza virus inactivation by UV-C

A quartz cuvette containing 2 ml of a 5.0 * 10E7 PFU/ml solution of M13 bacteriophage was positioned in the UV-C disinfection device next to a quartz cuvette containing 2 ml of a 5.0 * 10E7 PFU/ml solution of the PR8 H1N1 influenza virus. A disinfection cycle of 10 min was performed. The experiment was conducted 3 times, and the viral titers of the solutions were then measured through viral titration as described above.

### Foam and microphone contamination

A 10^11^PFU/ml bacteriophage M13 solution was sprayed on either the microphone foam (1×1 cm) or the microphone as a whole for 5 s using a Thermo Scientific Nalgene 2430-0200 aerosol spray bottle. The nozzle was positioned at a distance of 10 cm from the target objects and with an inclination angle of 45°. The bacteriophage solution was sprayed on the objects for 5 s, followed by air-drying of the objects for 1 h at room temperature. For the experiments we used the Shure SM58 foam and Sennheiser e835 microphones.

### Disinfection and recovery procedure of inoculated foam

The disinfection tests were performed in technical replicates and over all three times independently. 10 μl of a M13 suspension with 10^13^ PFU/ml was deposited evenly, by above mentioned spraying, on the surface of a 1 cm^2^ piece of foam (Shure SM58) cut from the microphone part, and the liquid was allowed to dry. To test how effectively the virus could be recollected from the materialthe dried foam was placed in a Falcon tube of 50 ml which was filled with 10 ml MilliQ water. The tube was placed for 15 h on a rotation device that allows for proper shaking and agitation of the foam, which freely moved inside the water to ensure maximum extraction. Foam samples that were to be disinfected were airdried after inoculation and mounted on a wooden cocktail stick to allow them to be placed in a similar position within the UV-C chamber as a microphone would be. Duplicate samples were irradiated for 10 min while the controls were left untreated. After irradiation, all samples were subjected to the extraction method for 15 h and the virus content was determined using the same method of inoculation and plating as described above. The difference in viral load between the disinfected sample and the respective control was calculated to assess the reduction in PFU.

### Disinfection and recovery procedure of inoculated microphones

The microphones, inoculated with M13 as described above, were placed in containers large enough for the microphones to be fully submerged in 400 ml MilliQ water. Next, the containers were placed for 15 h on a mixer performing a similar extraction as for the foam alone (Figure S2). After 15 h, the concentration of infectious virus particles in the extraction solution was determined through viral titration to assess the efficacy of UV-C treatment, following the same procedure as described above for assessment of disinfection of the foams.

## 3. Results

### UV-C device design and validation

A UV-C device was specifically tailored for the disinfection of microphones (Figure 2). The device is equipped with two Osram Puritec Germicidal lamps HNS 16W G5 with 254 nm as the maximum intensity wavelength. The device includes a drawer that slides out of a protective casing to provide access to the inner chamber that is equipped with a removable tray holding the microphones. Once closed, there is no UV-C radiation leakage outside the box. Further, the mode of operation is such that the only possible variation is the time of irradiation, which simplifies the operation of the device. While different microorganisms require different intensities of irradiation, such requirements can simply be met by lengthening the irradiation time as the process is cumulative. Thus, to achieve twice the irradiation intensity it will suffice to double the irradiation time.

**Figure 2.**
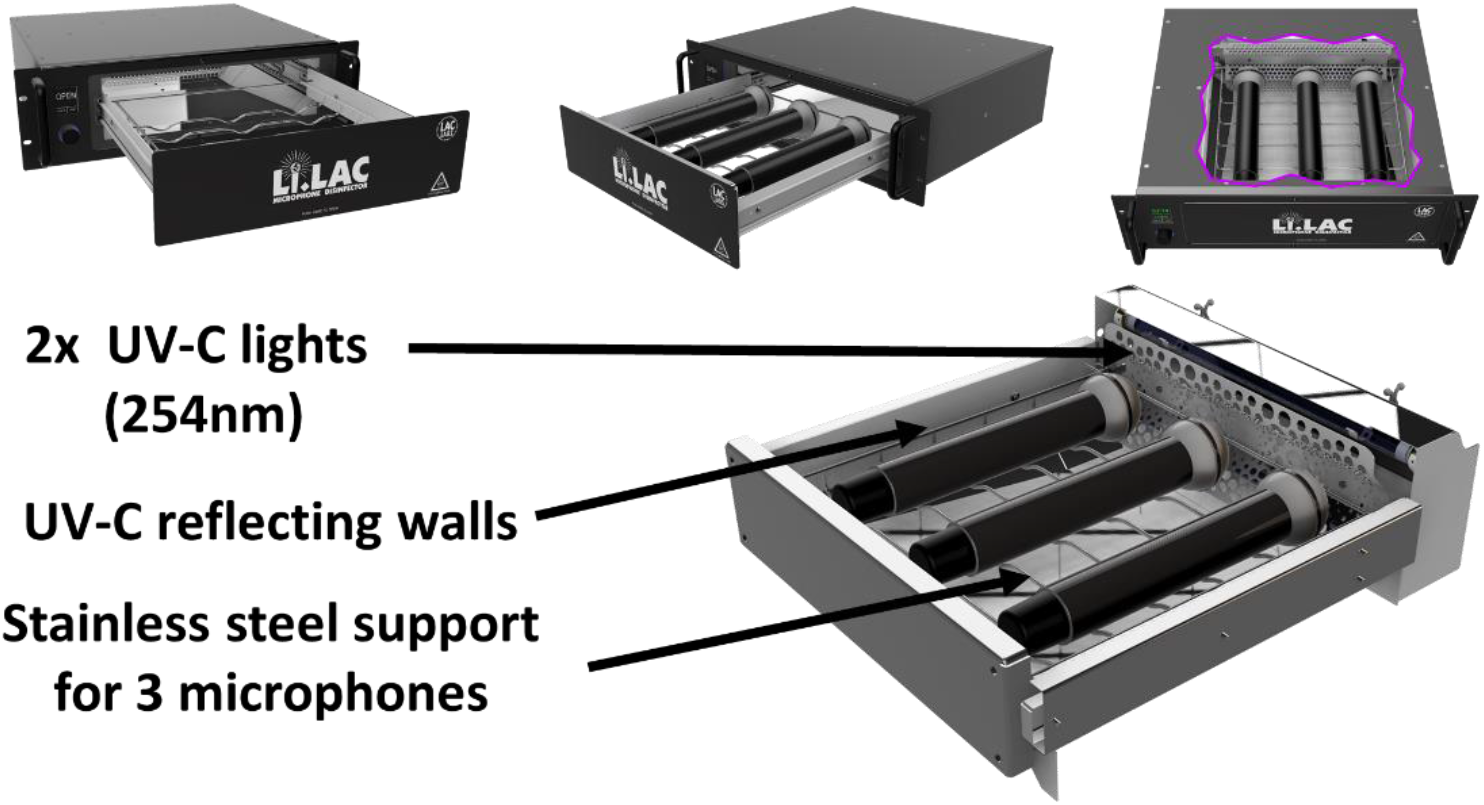
Basic design of the UV-C disinfection device that can contain up to three microphones at the same time. The microphone support slides into a shielded casing that contains two UV-C lamps. The efficiency of irradiation is enhanced by the usage of UV-C reflecting walls.

Microphones are complex pieces of equipment in terms of shape. Hence, careful design of their placement within the UV-C disinfection chamber is needed in order to assure that everywhere within the device, the irradiation intensity is high enough to effectively eliminate contaminating micro-organisms, while keeping the irradiation times short enough to remain practical. As is shown in Figure 2, the UV-C sources were placed at one end of the chamber and the walls of the inner chamber were made to reflect UV-C to minimize loss due to UV-C absorption. Consequently, the items that most effectively absorb the irradiation are the microphones of which up to three can be placed within the chamber for disinfection. The irradiance (μW/cm^2^) of the UV-C was measured at various locations within the device according to an external accredited agency (Opsytec Dr. Groebel GmbH) (**Supporting Information 1**) and it was shown to range from 523 to 7185 μW/cm^2^ depending on the position within the device. Different measurements at the same locations showed variations of 100-150 μW/cm^2^. Judged by the lowest measured irradiance (i.e. 373 μW/cm^2^, Supporting Information 1), the theoretical efficacy for eliminating the most common pathogens would be >99.9% upon 1 min irradiation (according to DIN 5031-10:2018-03). While inside the UV-C device the irradiance is high enough for effective disinfection, the measured UV-C values outside the device were below 9 μW/m^2^ (**Supporting Information 2**), which is far below the acceptable limits (max dose: 1042 μW/m^2^ for 8 hours continuously). Therefore, we conclude that the device is safe to use.

### Assessment of virus extractability

Microphones have complex shapes and are fabricated from various materials, such as plastics and metals, which are also located on places that are difficult to reach in terms of cleaning as well as possible exposure to the UV-C irradiation. One of the main parts that was foreseen as a potentially problematic area was the inner foam, which resides underneath a metal grill at the microphone head. The foam has a porous structure and, hence, aerosols or liquids that penetrate into the foam and dry out would possibly be protected/shielded from UV-C irradiation rendering the treatment less effective. Therefore, we started by investigating the disinfection of the foam, which also allowed us to set up the basic methodology.

For the assessment of the efficacy of UV-C treatment, we used the M13 bacteriophage as it offers a straightforward readout in terms of infectivity. As described in the methods section, 10 μl of a M13 suspension with 10^13^ PFU/ml was used to contaminate the foam (Shure SM58). It was envisioned that shaking and agitation of the foam in water would allow us to extract the virus from the foam. Different mixing times were investigated by taking aliquots at different time points and assessing the concentration of the infectious viral particles in the solutions through viral titration. The detected viral concentration was then compared with the concentration measured for a control sample in which 10 μl of M13 at 10^13^ PFU /ml were directly transferred to 10 ml MilliQ water, serving as the 100% recollectable concentration. The intention was not to have a method that extracts 100% of the applied viruses, but to have enough virus extracted to perform reliable disinfection measurements with a minimal number of dilution steps. It was found that after 2 h of mixing, about 77% of the virus could be recovered from the foam, while after 15 h this number increased to 81%.

### UV-C disinfection assessment

#### UV-C inactivation of M13 and influenza virus in solution

To get first insight into the efficacy of the device and to allow extrapolating our findings to pathogenic viruses, we placed quartz cuvettes containing bacteriophage M13 or PR8 H1N1 influenza virus in the device and simultaneously irradiated the liquid samples for 10 min. The initial concentration before irradiation was 5,0 * 10E^7^ PFU/ml for both viruses. In all the samples no active viruses were detectable upon irradiation indicating an efficacy of over 99.99%, which concurs with the theoretical calculation of irradiance and viral susceptibility towards UV-C. The results indicate that M13 is a suitable model virus for these tests.

#### Foam disinfection

To determine the UV-C disinfection efficacy, samples of foam of 1 cm^2^ were used. The samples were contaminated via a spraying device to mimic aerosol formation and deposition. The spraying device was placed at a distance and angle that allowed us to cover two foam samples with aerosols simultaneously during a short burst. One sample served as control while the other sample was treated with UV-C (Figure 3). For the spraying deposition, it was found that more liquid was deposited than in the initial recovery experiments, where 10 μl aliquots of M13 suspension were applied to the foam by pipetting. Therefore, the M13 concentration used for the contamination of foams by spraying was reduced to 10^11^ PFU/ml. The exact volume could not be determined however, extractability and comparison between control and sample maintained correct.

**Figure 3.**
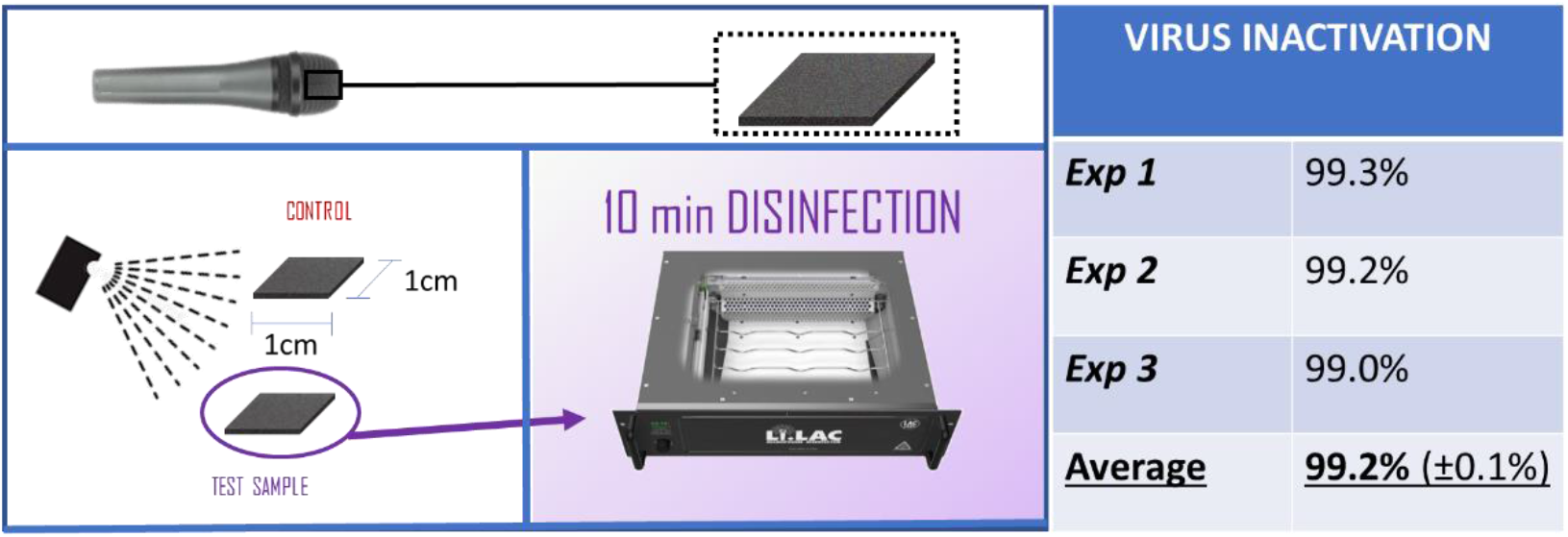
Representation of the spray deposition of virus onto a foam from the inside of the microphone, and representative results from disinfection experiments.

For the inoculated foam samples, the viruses were extracted and the viral loads of the disinfected sample and the respective control were compared to assess the reduction of PFU in percentage. Figure 3 shows reductions in the M13 titers of 99.3, 99.2, and 99.0% in the technical replicates. On average it was found that a 99.2% reduction in the viral titer was achieved by 10 min of UV-C irradiation. We regard this as an important finding, because the foam is considered to be the most inaccessible part of the microphone that may shield micro-organisms against UV-C due to its porosity. The efficacy of UV-C disinfection for 20 min was only marginally increased compared to disinfection for 10 min and, therefore, all further experiments involved 10 min UV-C disinfection.

### UV-C disinfection assessment of infected microphones

Since UV-C disinfection of microphone foams was very effective, we next evaluated the UV-C disinfection of microphones as a whole. To this end, the method of spraying and virus extraction was adapted to contaminate the whole microphone (Figure 4). Also in this case, two microphones were sprayed at the same time, receiving an equal dose of M13 bacteriophage, where one microphone served as the untreated control, while the other one was subjected to 10 min of UV-C irradiation. Subsequently, the microphones were extracted in a similar fashion as the foam, but using a larger container holding 400 ml of water. These experiments were performed three times independently with technical replicates.

**Figure 4.**
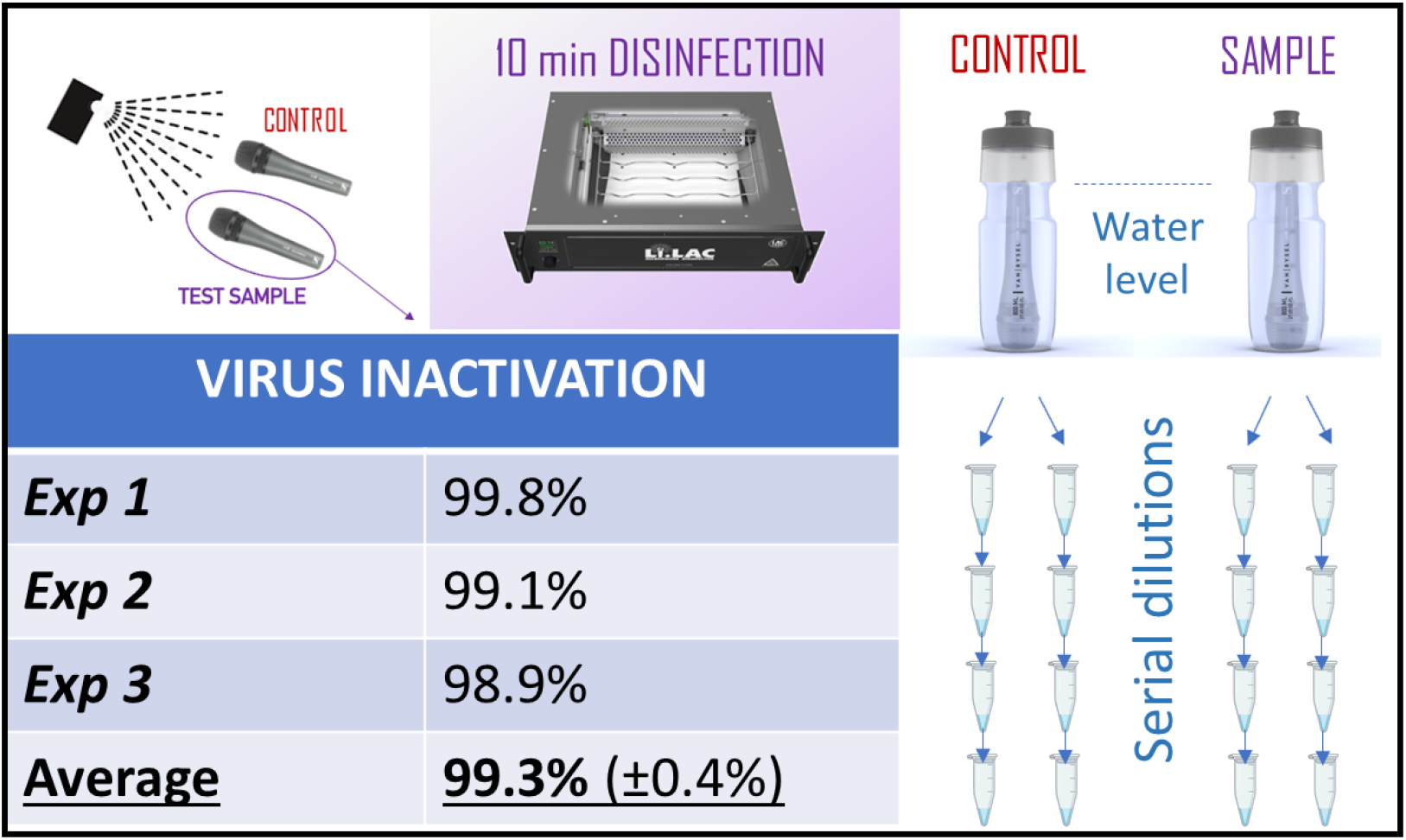
Spray deposition of bacteriophage M13 on complete microphones, which were subsequently subjected to disinfection using UV-C. Extraction of M13 was performed by full submersion of the microphones in water combined with shaking. The amounts of M13 extracted from infected and disinfected microphones were compared, and the total reduction of active virus was determined.

The PFU reduction on whole microphones upon UV-C irradiation was determined by comparing the control to the 10 min irradiated microphone, showing an average reduction in infectious virus particles of 99.8, 99.1, and 98.9% for the different technical replicates and an overall average reduction of 99.3%. The efficacy of the disinfection of the entire microphone was thus comparable to what was observed for disinfection of the foam.

## 4. Discussion

Disinfection of devices shared among individuals remains a vital precaution to prevent the spread of infections. The present COVID-19 pandemic as well as readily transmissible infections caused by various types of influenza illustrate the impact that infectious diseases can have in our daily lives, the well-being of both humans and animals, and the (socio)economic consequences [8,33]. Here, we present a facile disinfection device that can be used in events where microphones are predominantly used, which includes entertainment industry but also conferences and other social events, to prevent the transmission of infectious agents.

Since the M13 bacteriophage does not represent a threat to humans or animals, it was used here as a facile model to determine the efficacy of UV-C irradiation in the elimination of viral contaminations on microphones. Importantly, our verification experiments with PR8 H1N1 influenza virus showed that this virus was successfully eliminated by UV-C treatment with an efficacy that was indistinguishable from the efficacy at which the M13 bacteriophage was eradicated. However, it has to be noted that, for reasons of biosafety, these verification experiments were performed in solution, while the disinfection experiments with the foams and whole microphones involved dry surfaces contaminated with the M13 bacteriophage. Therefore, there might still be a difference in terms of the susceptibility of different viruses to UV-C treatment when present on dry surfaces.

Based on the theoretical considerations and the known susceptibility to UV-C (Influenza dose of 105 J/m^2^ according to DIN 5031-10:2018-03[34] but lower values have been measured [21]), a 1-min irradiation at 373 μW/cm^2^, the lowest irradiance value measured in the device, would be sufficient to achieve 99.9% of viral inactivation (According to [34]). Our experiments demonstrated a 99.3% reduction in the viral load upon 10-min UV-C disinfection of whole microphones. We consider this result impressive regarding the complex construction of a microphone which makes it very difficult to irradiate all deposited viral particles with equal efficiency.

While the present UV-C disinfection device was tested for microphones and the respective foams, the implications of this technology are much broader. In particular, disinfection strategies based on UV-C are of great interest in the medical field. The main challenge here is to apply standardized methodology, which is difficult to achieve as the conditions can vary greatly. For instance surfaces may be wet or dry, and microbial contaminants may be surface-bound or suspended, or strongly or weakly adherent. These factors could potentially cause differences in the overall efficacy of UV-C irradiation. Although, conditions may vary and responses can differ, here we show that when the controls are chosen properly, it is possible to approximate the UV-C treatment efficacy. Hence, not only the here-described device for disinfection can be regarded as a valuable tool, but also our established methodology for contaminating surfaces and re-extracting viral particles may contribute to standardizing UV-C-induced disinfection strategies.

## 5. Conclusions

Here we present a UV-C device capable of disinfecting virus-contaminated microphones in a short period of time, i.e. 10 min, with at least 99.3% efficacy. To this end, we successfully developed a viral inoculation and extraction protocol. The protocol involves aerosol production with a spraying device, which allows the pairwise contamination of objects, here microphones, with similar viral doses. The applied viral extraction method allows an approximation of the efficacy of viral inactivation by UV-C treatment. We are therefore confident that not only our UV-C disinfection device, but also the developed viral contamination and extraction approaches will positively contribute to the development of new avenues in infection prevention.

## Data Availability

All data produced in the present study are available upon reasonable request to the authors

## Supplementary Materials

The following supporting information can be downloaded at: www.mdpi.com/xxx/s1, Figure S1: title; Table S1: title; Video S1: title.

## Author Contributions

The contributions of the authors to this work are as follows: Conceptualization, T.H and P.R.; methodology, V.V., T.H., J.V-I., A.H., J.M.D., P.R.; formal analysis, V.V.; investigation, V.V.; data curation, V.V., J.V-I.; writing—original draft preparation, P.R.; writing—review and editing, V.V., T.H., J.V-I., A.H., J.M.D., P.R.; supervision, A.H., J.M.D., P.R.. All authors have read and agreed to the published version of the manuscript.

## Funding

This research was partially funded by LAC Labs GmbH, and the European Union’s Horizon 2020 research and innovation program under the Marie Skłodowska-Curie grant agreement No 713482 (ALERT Cofund).

## Data Availability Statement

Data is available on request from the authors.

## Conflicts of Interest

The authors declare the following conflict of interests: P. van Rijn also is cofounder, scientific advisor, and shareholder of BiomACS BV, a biomedical-oriented screening company. This research was partly funded by LAC Labs GmbH (T.H.) but did not influence the scientific merits or outcomes in any way. The other authors declare no conflicts of interest.

## Notes

### Competing Interest Statement

The authors declare the following conflict of interests: P. van Rijn also is co-founder, scientific advisor, and shareholder of BiomACS BV, a biomedical-oriented screening company. This re-search was partly funded by LAC Labs GmbH (T.H.) but did not influence the scientific merits or outcomes in any way. The other authors declare no conflicts of interest.

### Funding Statement

This research was partially funded by LAC Labs GmbH, and the European Unions Horizon 2020 research and innovation program under the Marie Skłodowska-Curie grant agreement No 713482 (ALERT Cofund)

