## Supplementary Information for "An efficient UV-C disinfection approach and biological assessment strategy for microphones"

**S1:** Expert data file external agency on radiance performance
**S2:** Expert data file external agency on radiance safety

**The official documents are receivable upon request due to language incompatibilities with MedRxiv. Please contact for the full Supporting Information**
